# Evaluating Student Use, Access, and Perceptions of a Campus Sexual Health Vending Machine

**DOI:** 10.64898/2026.07.03.26357232

**Authors:** Grace Sheng, Lisa Gualtieri

## Abstract

**Background:** Sexual health vending machines have emerged as a promising approach to expanding access to contraception and other sexual health products on college campuses. This study evaluated patterns of use, perceived accessibility, and student attitudes towards the sexual health vending machine at Tufts University.

**Methods:** This mixed-methods evaluation combined an online survey with vending machine refill data. Analyses included descriptive analysis, chi-square tests, and thematic analysis of open-ended responses.

**Results:** The survey was open from October to December 2025 and included 118 respondents. Self-reported use suggests students engage with the vending machine on an as-needed basis.

Users were significantly more likely to agree that it increased their likelihood of using a condom, compared with non-users. However, both groups expressed strong support for the vending machine, reporting that it improved access to sexual health resources, offered satisfactory product variety, and should be provided by the university. Although many students appreciated its visibility and role in reducing stigma, concerns about privacy and discretion were the most commonly reported reasons for non-use. Restock data indicated sustained year-round utilization, with expected decreases during vacation periods.

**Conclusion:** Overall, these findings suggest that sexual health vending machines represent a scalable strategy for expanding access to sexual health resources in university settings. Offering a unique combination of immediacy, affordability, and convenience, this low-barrier resource may also promote preventive sexual health behaviors across campus, even among non-users, and support broader efforts to prevent sexually transmitted infections and unintended pregnancy among young adults.

## Background

Sexually transmitted infections and unintended pregnancy continue to be public health priorities among adolescents and young adults, with national data underscoring persistent risk behaviors such as low or inconsistent contraceptive use [Szucs L.E. et al., 2019]. Given the potential lasting physical and psychosocial implications of STIs and unintended pregnancy, understanding patterns of contraception use in this population is an important area of public health research.

College students in particular experience high rates of inconsistent contraceptive use [Kusseling F.S. et al., 1995] and many encounter or perceive barriers to obtaining timely sexual and reproductive health products. Students frequently reported obstacles including cost, privacy concerns, inconvenient pharmacy or health center hours, embarrassment or stigma related to obtaining sexual health products, and lack of awareness of campus resources [Olson R. et al., 2024; Mozingo S.L. et al., 2023; Bersamin M. et al., 2017]. Such barriers may delay or prevent necessary contraceptive use, particularly in time-sensitive situations requiring emergency contraception. Ensuring rapid, convenient access to sexual and reproductive health resources is a key component of effective pregnancy and STI prevention efforts on college campuses [Bhochhibhoya S. et al., 2025].

In response, universities across the United States have begun placing vending machines on campus that dispense contraception and other sexual health products. Reports from 2023 showed that approximately 39 US universities in 17 states operated such vending machines [Luna I. 2023], a number that has grown to more than 150 campuses as of May 2026 [EC4EC, n.d.].

Tufts University, a private university in Medford, Massachusetts with 7,126 undergraduate and 6,473 graduate students [US News, n.d.], participates in this trend of increasing sexual health access in universities. The first sexual health vending machine was installed in October 2022 by the Center for Awareness, Resources, and Education (CARE) in collaboration with the Tufts student group Sex Health Reps, with the goal of encouraging safe sexual practices and reducing stigma surrounding sexual health [Anderson E., 2022]. The issue of accessibility, only exacerbated by the COVID-19 pandemic, has long been a central concern, so this appointment-free, grab-and-go resource was designed to provide students with a discreet, convenient, and reliable way to access sexual health products. Located on the ground floor of the campus center, external and internal condoms, dental dams, and lubricant are provided free of charge [AS&E Students, n.d.]. To ensure discretion, items can be obtained without a student ID.

### Objective

While early evidence suggests that these machines may increase access, reduce stigma, and provide critical after-hours availability at other universities [Knifton S. et al., 2024; Houlihan K. et al., 2026], a formal evaluation of the impact of Tufts’ machine had not been conducted, nor had an analysis of how factors such as student population, campus culture, machine location, and pricing influence use. Thus, this study aimed to explore student use of and attitudes toward the Tufts University Medford campus sexual health vending machine. Furthermore, most published studies only gathered survey data, but by combining refill counts with self-reported and other qualitative survey data, this study sought to gather a more comprehensive understanding of use, accessibility, and perceptions of the sexual health vending machine at Tufts.

## Methods

This mixed-methods study at Tufts University deployed an online survey and collected vending machine refill data.

An 18-question online survey was developed using Qualtrics to assess the impact of the sexual health vending machine. It included multiple-choice and open-ended questions about self-reported use of products, accessibility, barriers to use, and perceptions of the campus sexual health vending machine. Demographic data collected included year in school, gender identity, sexual identity/orientation, race/ethnicity, and place of residence during the academic year. The survey took approximately 5 minutes to complete. All questions required some response to complete and submit the survey, but participants were free to skip questions they felt uncomfortable answering by selecting “N/A”. Before university-wide distribution, the survey questions were piloted with 3 current Tufts University School of Medicine students who previously attended Tufts University as undergraduates. The Tufts Social, Behavioral, and Educational Research Institutional Review Board determined that IRB review and approval was not required.

The survey was open from October 30, 2025 to December 12, 2025. Participants were recruited with printed flyers containing a QR code to the survey, which were placed inside vending machine product envelopes. Advertisements were also posted on campus organization social media accounts, including Tufts Sex Health Reps, Planned Parenthood Action at Tufts, and the Tufts Sidechat forum. Students currently enrolled in classes at Tufts University who were 18 years or older were eligible to respond. The survey limited each student to one unique survey response. Survey responses contained no identifiable information and demographic characteristics were collected for the purpose of conducting statistical analyses. Anyone who completed the survey was eligible to win a $50 gift card by random draw, provided by CARE.

Survey data was analyzed using StataSE. Multiple-choice questions were analyzed using descriptive statistics, including count and frequencies. Demographic data were analyzed using a Chi-square test to assess for statistically significant differences between demographic subgroups. Statistical significance was defined as a *p* value less than or equal to 0.05. We also conducted thematic analysis of open-ended questions to identify patterns emerging from responses.

Machine refill data was collected from January 14, 2025 to December 5, 2025. Counts of products dispensed were extracted from restocking data provided by CARE and contained no identifying information.

## Results

### Demographics

A total of 118 students completed the survey. Participants were primarily undergraduate students (91.5%), composed of 16.4% freshmen, 38.8% sophomores, 22.4% juniors, and 15.5% seniors. The sample was predominantly female (77.4%) and included a diversity of sexual identities, with just over half identifying as heterosexual (53.0%). Approximately half of respondents identified as White (51.8%), followed by Asian (22.8%), Hispanic/Latino (12.3%), and multiracial (8.8%). Most participants resided on campus (62.1%) or off campus in the surrounding area (36.2%).

These data are presented in detail in Table 1.

**Table 1.** Participant demographic characteristics. Demographic characteristics of participants who completed the sexual health vending machine survey, including academic status, gender identity, sexual orientation, race/ethnicity, and residence status (n=114–116, depending on item response).

| Characteristic | Total |
| --- | --- |
| Year in school (n=116) |  |
| Freshman (1st year) | 19 (16.4%) |
| Sophomore (2nd year) | 45 (38.8%) |
| Junior (3rd year) | 26 (22.4%) |
| Senior (4th year) | 18 (15.5%) |
| Graduate student | 7 (6.0%) |
| Other | 1 (0.9%) |
| Gender identity (n=115) |  |
| Male | 21 (18.3%) |
| Female | 89 (77.4%) |
| Non-binary | 5 (4.4%) |
| Sexual identity / orientation (n=115) |  |
| Heterosexual | 61 (53.0%) |
| Bisexual | 30 (26.1%) |
| Gay / Lesbian | 11 (9.6%) |
| Pansexual | 6 (5.2%) |
| Asexual | 1 (0.9%) |
| Other | 6 (5.2%) |
| Race/ethnicity (n=114) |  |
| White | 59 (51.8%) |
| Asian | 26 (22.8%) |
| Hispanic / Latino | 14 (12.3%) |
| Black / African American | 2 (1.8%) |
| Native Hawaiian / Pacific Islander | 1 (0.9%) |
| Middle Eastern / North African | 2 (1.8%) |
| Multiracial / Mixed | 10 (8.8%) |
| Place of residence (n=116) |  |
| On campus dorm / apartment | 72 (62.1%) |
| Off campus - Surrounding area | 42 (36.2%) |
| Off campus - Commuter | 2 (1.7%) |

### Vending Machine Use

Of the 118 respondents, almost all (98.3%) were aware of the vending machine prior to taking the survey. Many of the respondents learned about the vending machine through campus promotion (61.4%), while others heard about it through friends (24.6%), social media (21.1%), seeing it in the campus center (19.3%), or through resident assistants (4.4%).

Most participants (66.7%) reported not having used the vending machine in the past 30 days. An additional 23.1% reported using it once, while 10.2% used it two times or more. Among vending machine users, 70.0% obtained condoms (external or internal), 40.0% obtained lubricant, 7.5% obtained dental dams, and 40.0% obtained other products. When asked about the circumstances for obtaining vending machine products, students most commonly reported “to keep on hand just in case” (n=40), 25 “in response to an unplanned need,” and 20 “right before sexual activity.” Vending machine use did not differ significantly between those that lived on and off campus (χ^2^, p=0.318).

### Accessibility and Barriers

When asked if they have ever wanted to use the machine but decided not to, 38.1% of respondents answered “yes.” Of those, the most cited reason was visibility/privacy concerns (81.8%), followed by not knowing how to use the machine (10.9%), cost (9.1%), and the desired product being out of stock (1.8%).

Most respondents reported they would obtain sexual health products from a pharmacy (83.2%), campus health center (31.9%), or resident assistant (17.7%) if the vending machine was not available on campus. Other answers included online (12.4%), from friends (7.1%), or other (1.8%).

Over half of respondents felt that having the vending machine on campus increased their likelihood of using a condom, with 28.0% strongly agreeing and 31.4% somewhat agreeing. 22.9% neither agreed nor disagreed, 0.9% somewhat disagreed, 3.4% strongly disagreed, and the remaining 13.6% selected ‘N/A.’ Furthermore, students who reported using the vending machine in the past 30 days were significantly more likely than non-users to agree that its presence increased their likelihood of using a condom (χ ^2^, p=0.025).

### Perceptions and Attitudes

There was a very strong consensus among respondents that the vending machine makes it easier to obtain sexual health products, with 77.6% strongly agreeing, 20.7% somewhat agreeing, only 1.7% neither agreeing nor disagreeing, and no respondents disagreeing. This did not differ significantly by vending machine use (χ^2^, p=0.307), with 100% of users and 97.4% of non-users endorsing the statement.

Most participants were either very satisfied (64.4%) or somewhat satisfied (25.0%) with the variety of products offered in the vending machine, with only 9.6% neither satisfied nor dissatisfied and 1.0% somewhat dissatisfied. This did not differ by vending machine use (χ^2^, p = 0.220), with 92.3% of users and 87.7% of non-users reporting feeling satisfied. This also did not differ by year in school (χ^2^, p=0.405), gender identity (χ^2^, p=0.872), sexual identity/orientation (χ^2^, p=0.854), race/ethnicity (χ^2^, p=0.980), or place of residence (χ^2^, p=0.760).

Nearly all respondents felt Tufts should be providing sexual health resources in this way, with 88.1% indicating they felt it was very important and 11.0% somewhat important. Only 1 respondent selected ‘N/A’ and no respondents rated the approach as unimportant; therefore, inferential comparisons by vending machine use were not conducted.

### Participant Feedback and Suggestions

In response to an open-ended question about additional products, participants most commonly suggested different condom sizes (n=6), greater lubricant variety (n=4), and menstrual products (n=4); less frequently mentioned items included spermicide, diaphragms, sex toy wash, and drink-sticker lids (each n=1). Regarding another open-ended question about experiences with the vending machine, participants most frequently commented on discretion and cost. Some expressed appreciation for reduced cost (n=8), convenience (n=4), and discretion (n=4), while others raised concerns about insufficient discretion (n=11); a small number described the machine as either easy or confusing to use (each n=1). Several respondents also described the vending machine as contributing to reduced stigma and increased visibility of sexual health resources on campus, noting that “having it as an option and seeing others use it has made it less stigmatized and accessible for people” and that they appreciated it as “a resource and visible marker of sex positivity/education on campus.”

### Vending Machine Restock Data

Monthly restock volumes varied but indicated consistent utilization of the vending machine across the calendar year, with expected decreases corresponding to when campus is less populated (summer and winter academic breaks). Condoms were restocked in greater quantity and more frequently than lubricant or dental dams (Figure 1).

**Figure 1.**
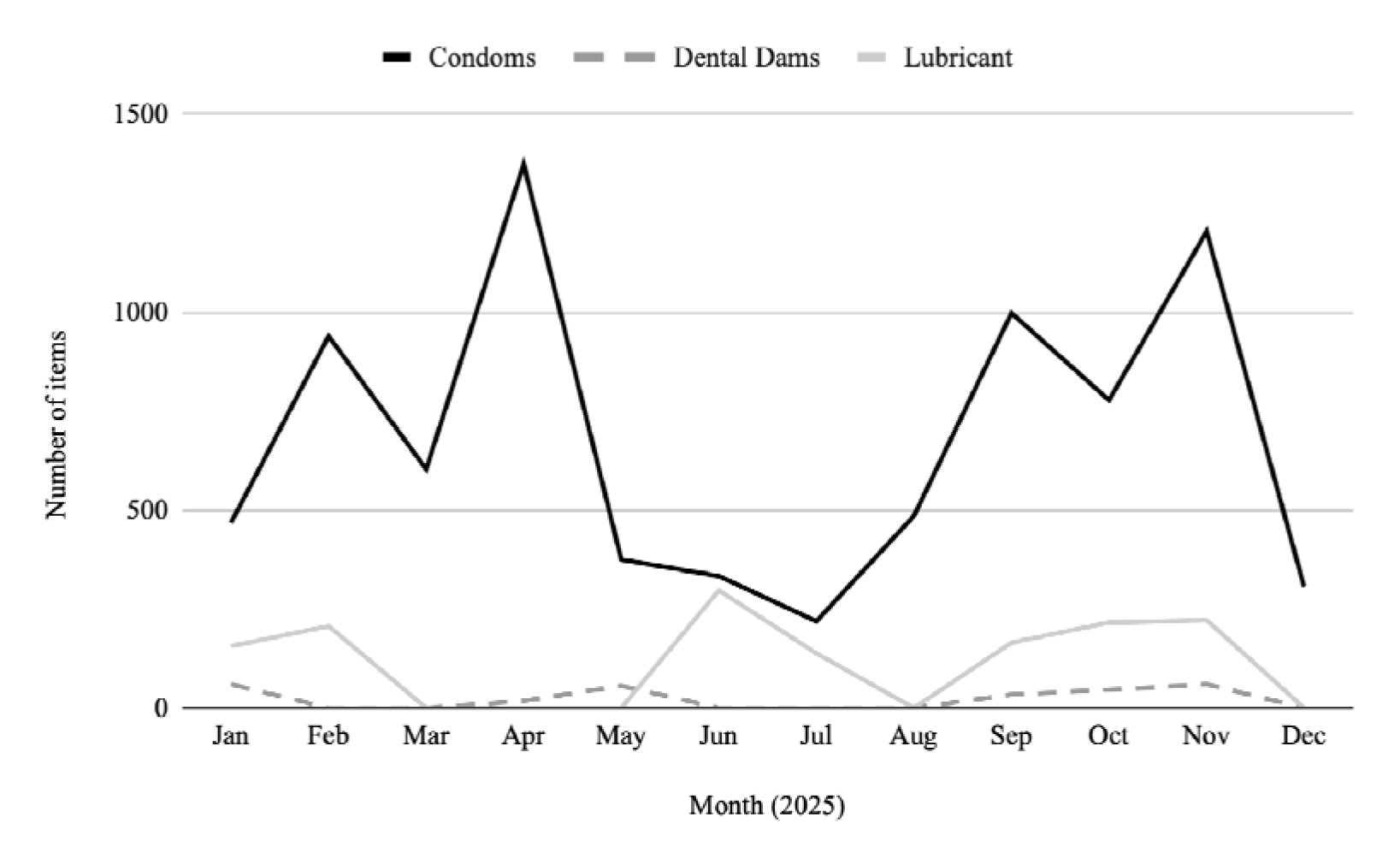
Monthly restocking of sexual health vending machine products. Monthly counts of restocked condoms (external and internal), dental dams, and lubricant in the campus sexual health vending machine over a 12-month period.

## Discussion

In this study, the majority of students reported not having used the sexual health vending machine in the past 30 days. However, perceptions of the resource were overwhelmingly positive and outweighed the proportion of recent vending machine users. Over 90% of participants agreed that the vending machine made sexual health products easier to obtain, were satisfied with the variety of products offered, and felt that the university should provide sexual health resources in this manner. This distinction suggests that the perceived value of the vending machine extends beyond direct utilization, with recognition of its role as a campus sexual health resource benefiting the broader student body.

Although most respondents reported no use of the vending machine in the past 30 days, this pattern likely reflects the inherently intermittent nature of need for sexual health products rather than lack of utility. Consistent with this, students most commonly reported obtaining products to keep on hand “just in case,” in response to an unplanned need, or immediately before sexual activity. Restock data demonstrates very consistent replenishment throughout the year, apart from expected declines during vacation periods, supporting the idea that the vending machine serves as a reliable resource when needed. Furthermore, restock data from the survey period (October 30 to December 12, 2025) show substantially higher volumes than self-reported use captured by the survey—with 1,509 condoms, 60 dental dams, and 222 units of lubricant replenished in November and December—suggesting that survey responses largely underestimate overall utilization of the vending machine.

Most participants reported being very satisfied or somewhat satisfied with the variety of products offered in the vending machine, with no significant differences between recent users and non-users or among any demographic characteristic. While these findings indicate that the current product selection is adequately serving a diverse range of students, participant suggestions for additional products—including different condom sizes, expanded lubricant options, and menstrual products—point to opportunities for further aligning the vending machine with student needs. Incorporating these items may enhance the relevance and inclusivity of the resource.

Over half of participants indicated that having the vending machine on campus increased their likelihood of using a condom, with students who reported recent vending machine use significantly more likely to endorse this effect than non-users. This association is consistent with other condom distribution studies demonstrating that access to low-barrier, readily available resources may support safer sexual behaviors among those who engage with them [Francis D.B. et al., 2016; Wang T. et al., 2018]. Notably, this proportion exceeded the number of students who had used the vending machine in the past 30 days, suggesting that the machine’s impact may extend beyond direct use. This finding raises the possibility of a broader ripple effect, in which the presence of visible sexual health resources contributes to shaping attitudes and intentions related to contraception use at a larger population level, even among non-users [Eisenberg M.E. et al., 2013; Cassidy C. et al., 2018; Sanyaolu et al., 2026]. Together with the high levels of perceived utility and satisfaction observed among respondents, this supports the idea that campus-based sexual health interventions can function as both access points and signals that reinforce preventative health behaviors.

Students’ strong agreement that the vending machine improves access further highlights its role as an important public health intervention with broad reach across the campus population. Several participants explicitly described the machine as reducing stigma and increasing the normalization of sexual health resource use, noting that “having it as an option and seeing others use it has made it less stigmatized and accessible for people” and that they appreciated it as “a resource and visible marker of sex positivity/education on campus.” In this way, the vending machine may function not only as a distribution mechanism but also as a visible cue that sexual health is a routine and accepted aspect of overall health and well-being, contributing to the destigmatization and normalization of preventive behaviors. However, this same visibility was also identified as a barrier to use, with some students expressing concern about being seen obtaining products, thus highlighting a tension between promoting accessible sexual health resources and preserving individual privacy.

Most respondents indicated that they would obtain sexual health products from a local pharmacy if the vending machine was unavailable. Despite the availability of online purchasing, relatively few chose this as a preferred alternative, suggesting discretion alone may not fully explain use patterns. Instead, the vending machine appears to offer a unique combination of immediacy, affordability, and convenience that is not fully replicated by existing alternatives.

These benefits include extended operating hours, elimination of the need for appointments, and free or lower-cost access to essential products, which may be particularly important for students with limited flexibility to access healthcare services during standard business hours. This is further supported by the finding that vending machine use did not differ by residence. Even among off-campus students, who may have greater access to pharmacies and other retail sources, utilization of the vending machine remained comparable to the on-campus group, suggesting that the resource provides benefits beyond geographic convenience alone. The vending machine may also better serve students who prefer to avoid in-person healthcare encounters due to discomfort, stigma, or other barriers associated with traditional clinical settings.

This study has several limitations. Survey responses were self-reported and may be subject to recall or social desirability bias. While the sample reflects a diverse student body, recruitment relied on convenience sampling at a single institution and may not be representative of the broader US college student population. As such, the generalizability of these findings to institutions with different demographic profiles, campus cultures, or resource availability may be limited. Additionally, vending machine restock data reflect aggregate replenishment rather than individual use patterns and cannot be directly linked to survey respondents.

Despite these limitations, this study contributes to a growing body of evidence supporting sexual health vending machines as valuable campus resources that increase access to sexual health products while fostering awareness and reducing stigma surrounding their use. Future research should examine longitudinal use patterns and impacts on sexual health outcomes across diverse campus settings to better understand the generalizability of these findings.

## Conclusion

This study provides evidence on the utilization and perceived value of a campus sexual health vending machine. Patterns of self-reported use suggest that individuals engage with the vending machine on an as-needed basis, consistent with the inherently situational nature of sexual health needs. However, restock data demonstrates steady and substantial utilization throughout the study period and over time. Together, these results indicate the vending machine serves as a valuable and accessible complement to existing campus health services and contributes to the normalization of sexual health resource use. Considering that visibility of the machine functioned as both a facilitator and a barrier to use, future initiatives might consider expanding access through additional vending machine locations, including more private settings that would allow students greater discretion while maintaining the visibility and awareness benefits of a centrally located machine. Overall, these findings support campus sexual health vending machines as a scalable public health strategy to reduce barriers to preventive care, expand access to essential sexual health resources, and strengthen health-promoting norms among young adults in higher education settings.

## Data Availability

Data in this study is not available due to privacy and confidentiality concerns.

## List of Abbreviations

(CARE): Center for Awareness, Resources, and Education

## Declarations

### Conflict of Interest

The authors declare that the research was conducted in the absence of any commercial or financial relationships that could be construed as a potential conflict of interest.

### Authors Contributions

GS conceived and designed the study, developed and administered the survey, collected and analyzed the data, interpreted the findings, and drafted the manuscript. LG provided study supervision, contributed to interpretation of the findings, and critically revised the manuscript. All authors read and approved the final manuscript.

### Funding

Funding for open access was provided by Tufts University Hirsh Health Sciences Library’s Open Access Fund. Otherwise, this research did not receive any specific grant from funding agencies in the public, commercial, or not-for-profit sectors.

## Acknowledgements

The authors would like to thank CARE for their valuable support of this project, including assistance with participant recruitment, funding of participant gift card incentives, and sharing of vending machine refill data. We also thank Drs. Ryan Main and Isaac Baek for their thoughtful review of the manuscript and valuable feedback during its development.

## Notes

### Competing Interest Statement

The authors have declared no competing interest.

### Author Declarations

Ethics Committee/IRB of Tufts University School of Medicine waived ethical approval for this work.

